# Knowledge, Practice, And Associated Factors Of Nurses Related To Pressure Ulcer Prevention In Selected Hospitals In Southern Ethiopia”, 2024

**DOI:** 10.1101/2025.03.04.25323368

**Authors:** Yohannes Tefera Ayane, Matusala Mamo Alembo, Ephrem Geja

## Abstract

**Background:** pressure ulcer is skin and tissue damage from pressure and friction, risky for immobile patients but preventable with early care. It harms health, causes discomfort and immobility, delay recovery, and nurses working in clinical settings plays important role in identifying patients at risk and administering preventative care.

**Objectives:** to assess nurses’ knowledge and practice towards pressure ulcer prevention and its associated factors in selected hospitals in southern Ethiopia, 2024.

**Method:** A cross-sectional study design was conducted in selected hospitals in Southern Ethiopia Regional State from May to June 2024. Data were collected using a structured self-administered paper questionnaire from 225 participants using a simple random sampling procedure, and the collected data were checked for their completeness with kobotool box and exported to SPSS version 26 for analysis.

**Result:** In this study, 139 (61.9%) of nurses have good knowledge and 137 (60.9%) of nurses have good practices. Having good knowledge [AOR = 6.940(3.527-13.656)], use of pressure ulcer prevention guidelines [AOR 3.3574(1.317-8.539)], formal training on pressure ulcer prevention [AOR 2.253(1.044-4.862)] and nursing leadership [AOR 2.246(1.064-4.742)] were associated with pressure ulcer prevention practices of nurses. While experience [AOR 3.52 (1.343-9.244)], marital status [AOR 0.414(0.219-0.780)], use of pressure sore guidelines [AOR 2.695(1.209-6.008)] and formal training [AOR 2.968(1.508-5.843)] were associated with pressure ulcer prevention knowledge of nurses.

**Conclusion:** The overall level of pressure ulcer prevention knowledge and practice of nurses were good. Knowledge, use of pressure ulcer prevention guideline, formal training on pressure ulcer prevention, nursing leadership, experience, and marital status were statistically significant factors for pressure ulcer prevention knowledge and practices. Health managers should offer ongoing professional development and updated guidelines to hospitals to enhance nurses’ knowledge and practices in preventing pressure ulcers.

## 1. INTRODUCTION

A pressure ulcer (PU) represents a localized injury that generally develops beneath medical devices or equipment, impacting the skin and/or tissue over bony prominences. This condition arises from prolonged pressure or a combination of pressure and shear forces (Zhang et al., 2021). The primary factors contributing to the development of PUs are external elements such as high pressure and shear. However, intrinsic factors can also play a role in the emergence of pressure ulcers, including limited mobility, inadequate nutrition, advanced age, conditions necessitating acute care, and spinal cord injuries (Guerrero et al., 2023). Individuals with spinal cord injuries, the elderly, and those who are sedated due to trauma or surgical procedures are at an increased risk of developing a pressure injury (PI), as they often remain in bed for extended periods and have fewer chances to reposition themselves (Berihu et al., 2020). To effectively prevent and manage pressure ulcers (PU), it is essential to evaluate the patient, their skin condition, and their individual risk factors (Guest et al., 2018). Nurses are the primary caregivers for bedridden and critically ill patients, who are particularly susceptible to pressure sores (Muhammad et al., 2017). Pressure ulcers can occur in any healthcare setting, with estimates indicating that 57% to 60% originate in hospitals, particularly within the first two weeks following a patient’s admission (Hospital, Zone and Birhan, 2022). Research conducted in Ethiopia highlights that pressure ulcers are a significant health concern among hospitalized patients, with a prevalence rate of 16.8% (Gedamu, Hailu and Amano, 2014; Hospital, Zone and Birhan, 2022). It is believed that improving nursing staff’s understanding and application of pressure ulcer prevention strategies is vital (Gunningberg et al., 2015). While the prevention of pressure ulcers is a shared responsibility across multiple disciplines, nurses typically assume a crucial role, which is considered a fundamental component of nursing care in high-income countries. Therefore, all nurses should prioritize prevention efforts.

## 2. Material and Methods

### 2.1 Study design, area and period

Institutional based cross sectional study design was conducted at selected public hospitals in Southern Ethiopia Regional State from May to June 2024. The selected public hospitals were Dilla University Referral Hospital (DURH), ArbaMinch General Hospital (AMGH) and Jinka General Hospital (JGH). DURH is located at Gedeo zone in Dilla town and is found on the main road from Addis Ababa to Nairobi, approximately 360 km from Addis Ababa, and 154 km from Wolaita Sodo. There are 142 nurses in DURH. The hospital has more than 2 million people of catchment area that lives in Dilla town, surrounding zones of southern Ethiopia, Sidama and Oromia region.

Whereas ArbaMinch General Hospital AMGH is located in southern, Ethiopia it is one of general hospitals in southern regional state of Ethiopia of Gamo Zone. It is situated in ArbaMinch Town, the capital of Gamo Zone. The town is found about 505 km South of Addis Ababa and 122 km from Wolaita Sodo, the capital of the Southern Ethiopia. There are 154 of total nurses in AMGH.

Jinka General Hospital is one general hospital found in southern Ethiopia regional state and which was established in 1991.The hospital has 120 beds and gives inpatient and outpatient services in its general outpatient department. Jinka town is the capital of South Omo zone located 691 km away from Addis Ababa and 267 km away from Wolaita sodo of southern Ethiopia Regional State. There are 137 nurses in Jinka General Hospital.

### 2.2 Population

#### 2.2.1 Source population

All Nurses working in southern Ethiopia hospitals.

#### 2.2.2 Study population

Nurses who were working in selected hospitals fulfilling the inclusion criteria.

#### 2.2.3 Study unit

All individual nurses who were selected for study.

### 2.3 Inclusion and exclusion criteria

#### 2.3.1 Inclusion criteria

All nurses who were permanent recruit working in selected hospitals.

#### 2.3.2 Exclusion criteria

Nurses who were being on annual, maternal, sick leave during data collection time.

#### 2.3.3 Sampling size determination and sampling techniques

#### 2.3.4 Sampling size determination

The sample size was determined by using a single population proportion formula with using a proportion of (54.4%) which was derived from a study conducted in Gondar University Hospital, Northwest Ethiopia (Tesfa Mengist *et al*., 2022).Confidence level at 95% and marginal error of 5%. The sample size was calculated as follows: the total calculated samples size was 381, and adding a 10% non-response rate and the final sample size was 419.

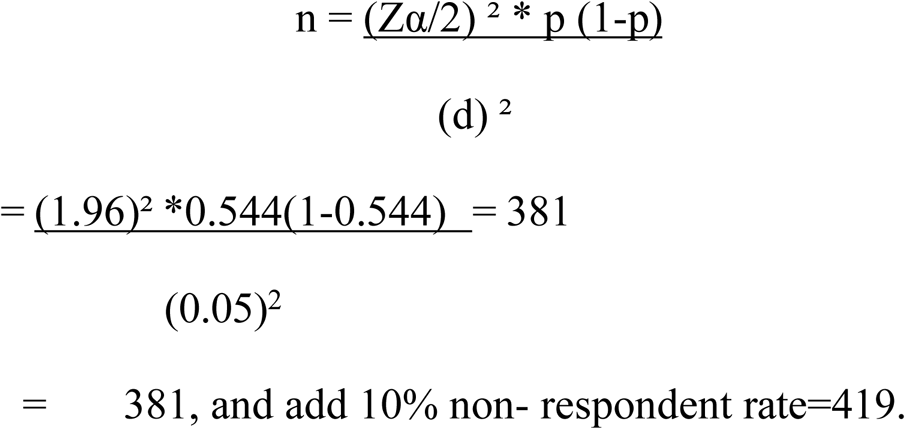

Since the total population is 433, the sample size was determined using correction formula, and adding a 10% non-response rate and the final sample size was 235.

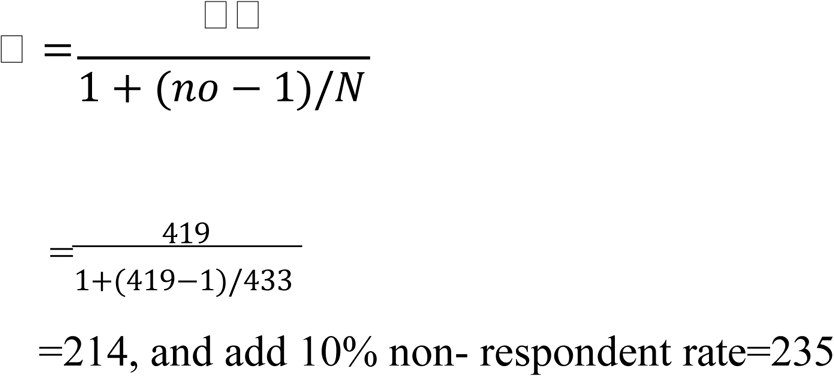

#### 2.3.5 Sampling techniques

The list or frame of nurses working in selected hospitals was obtained from the monthly work schedule prepared by the head nurse of each ward. There were six public general and refe hospitals in Southern Ethiopia Regional State and from those three (DURH, ArbaMinch, and Jinka General) hospitals were selected using simple random sampling, lottery method. A total of 433 nurses working in three selected hospitals selected: 84 nurses from a total of 154 nurses working in ArbaMinch general hospital; 77 nurses from 142 working in Dilla University referral hospital; 74 from a total of 137 working in Jinka general hospital; were selected, with a final number of 235 study subjects; nurses was selected proportionally using simple random sampling method from each hospital.

**Fig 1.**
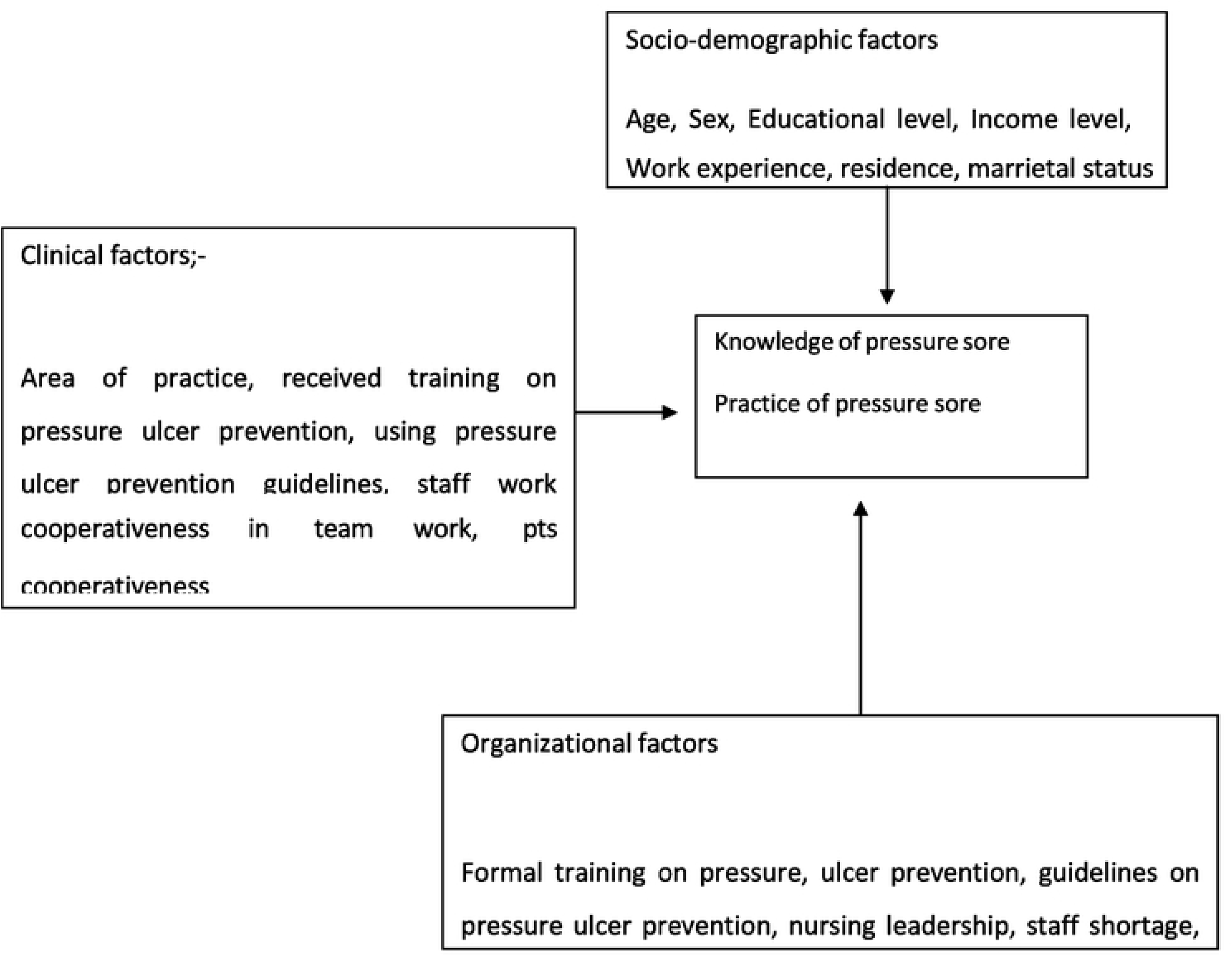
conceptual framework of knowledge, practice and associated factors of PUP among nurses in selected hospitals in southern Ethiopia (Nuru *et al.,* 2015; Tesfa Mengist *et al.,* 2022).

**Fig 2.**
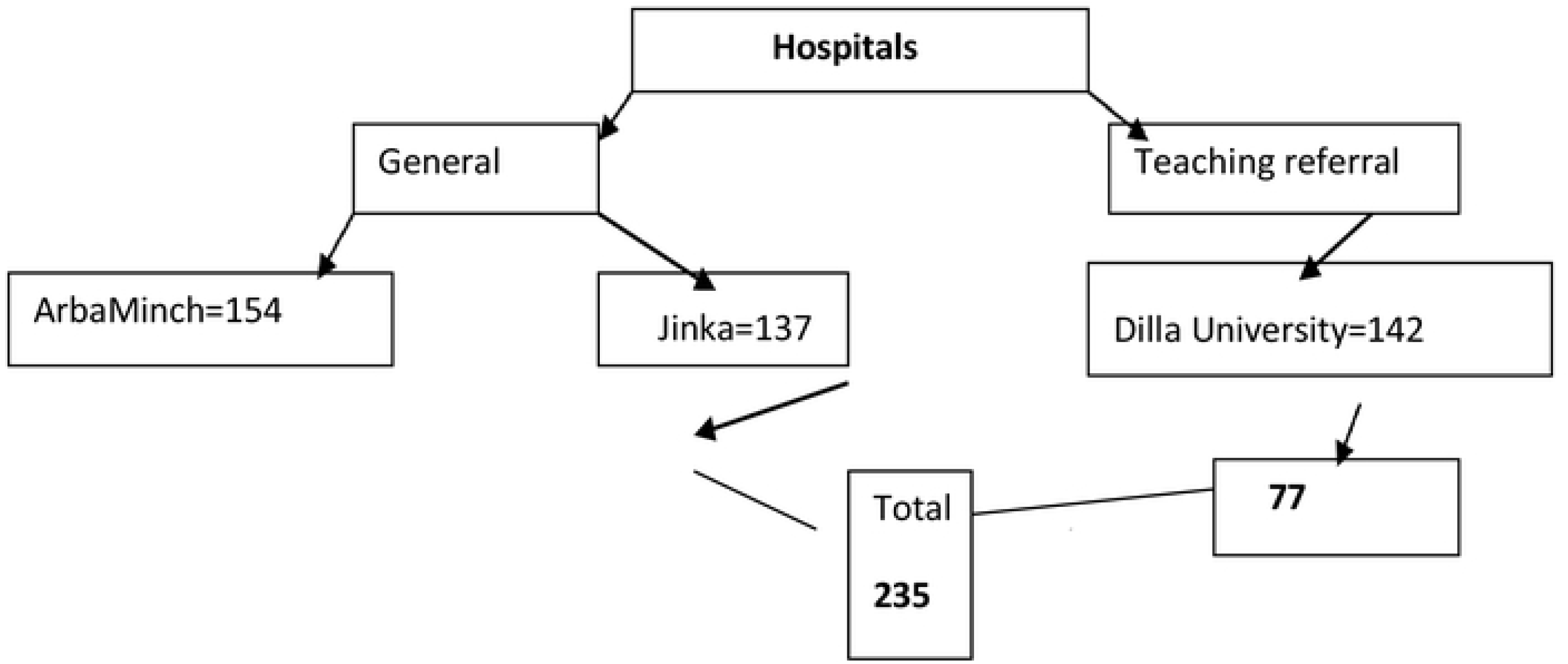
diagrain1natic representation of sa1npling technique

#### 2.3.6 Study variables

#### 2.3.7 Dependent variables

Practice of nurses towards pressure ulcer prevention

Knowledge of nurses towards pressure ulcer prevention

#### 2.3.8 Independent variables

Socio-demographic factors like, age, sex, income level, work experience, level of education

**Organizational factors**; - Formal training on pressure ulcer prevention (PUP), guidelines on PUP, nursing leadership, staff shortages, inadequate facilities and equipment, and type and level of institution.

**Clinical factors** like; - heavy work load, job satisfaction, lack of time, patient factor (uncooperative), area of practice, using PUP guideline, received training on PUP, unproportioned nurse; patient ratio.

#### 2.3.9 Operational definitions

##### Knowledge

Good knowledge: Nurses, who scored above the mean score of the knowledge questions towards pressure ulcer (Tesfa Mengist *et al*., 2022).

Poor knowledge: Nurses, who scored mean and above the mean of the knowledge questions towards pressure ulcer (Tesfa Mengist *et al*., 2022).

##### Practice

Good practice: Nurse who scored greater or equal to the mean of practice-related questions(Muhammad *et al*., 2017).

Poor practice: Nurse who scored less than the mean of practice-related questions (Muhammad *et al*., 2017).

## 3 Data collection and processing

### 3.1 Data collection tools and procedure

The structured questioner for knowledge and practice of pressure ulcer was adapted from study conducted in Gondar university hospital and Afghanistan (Muhammad *et al*., 2017; Tesfa Mengist *et al*., 2022),with the internal consistency reliability (Cronbach’s α) correlation coefficient of 0.88. But, socio-demographic items modified according the socio-cultural context of the community where study was done. Six BSC nurses and three Public Health Officers collected the data with close supervision. The questionnaire tool has five sections: socio-demographic part (sex, age, work experience, marital status, educational status, monthly salary), clinical factors (received training, use of guidelines, unproportionate patient nurse ratio, staff workload, staff cooperativeness, patient cooperativeness, and area of practice) and organizational factors (formal training on pressure ulcer prevention, staff shortage, inadequate guideline on pup, inadequate facility equipment, type of institution, and nursing leadership). Pressure Ulcer Prevention Knowledge and Practices Assessment Instrument were used for dependent variables. The instrument consists of 22 knowledge and 10 practices based questions.

Data quality was controlled by giving trainings and appropriate supervisions for data collectors. The overall supervision was carried out by the principal investigator. A pre-test was conducted using 10 % of the questionnaire on nurses who were working in Wolaita Sodo University Comprehensive Specialized Hospital. An appropriate modification was made after analyzing the pretest result before the actual data collection. A total of 235 study participants was selected using simple random sampling technique, where each member of the population has equal chance of being selected for final survey. Request for consent from nurses prior the collection. Continuous follow-up and supervision was given by supervisors throughout the data collection period.

### 3.2 Data processing, analysis and presentation

The questionnaires collected by kobotool.box were checked for completeness and exported to SPSS version 26 for further analysis. Descriptive statistics like texts, tables, graphs, and frequency was used to present participants’ characteristics. Both bivariate and multivariate logistic regression models were used to identify associated factors. Odds Ratios and their 95 % Confidence Intervals were computed and variables with p - value less than 0.05 was considered as significantly associated with the outcome variable.

#### 3.2.1 Data quality assurance

To maintain the quality of the data, the data collectors were receive training on data collection procedure. The questioner was designed carefully, and the English version was translated to Amharic version, and back to English then used for the actual data collection. Before the actual data collection, the questioner was checked for clarity and comprehensiveness. The instrument was tested by taking 10% of the sample size to check the validity and reliability of the tool, and modification was done accordingly. Then the collected data was cross checked for completeness and consistency each day by the principal investigators.

#### 3.2.2 Ethical clearance

Ethical approval was obtained from institutional Review Board (IRB) of HUCMHS with ethical approval number (Ref. No: IRB/154/16 on 20/03/2024. An official letter of co-operation was also written to concerned body from Hawassa University Nursing department and a letter was submitted to selected hospitals in Southern Ethiopia Regional State to get permission for conducting the study on population. The purpose of the study was explained to the study participants and written consent was secured before data collection is commenced and confidentiality of the information was be ensured. The start and end date of the recruitment period of this study was 20 march 2024 to 19 march 2025

## 4 Results

### 4.1 Socio demographic characteristics of the nurses

A total of 235 professional nurses were invited to participate in the study, and the response rate was 95.7% of which the majority of the respondents 130 (57.7%) were males. Moreover, the age of the participants ranged from 20 to 30 years, (74.2%) (n= 167) with a mean age of 26.45 ± 3.316 years. More than half (56%) were single whereas 44% of them were married. Regarding the educational status, the majority of them 85.8 (n=193) were first-degree holders, while 14.2% (n=32) were second degree holders.

Furthermore 12% (27) of these respondents had 1-2 years’ experience, about 57.8 %(130) of the respondents had 2-5 years’ experience and about 30.2 %(68) of the respondents had more than 5 years’ experience. Among the participants, 82.2 %(n=185) had a monthly income ranging from 6193 to 8017 Ethiopian birr and 17.8% (n=40) had a monthly income above 8017 Ethiopian birr. (Table1).

**Table 1.**
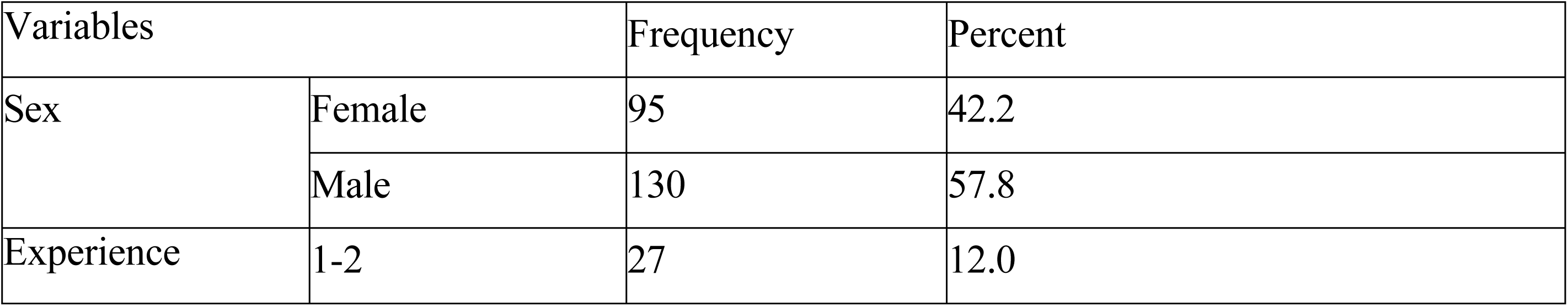
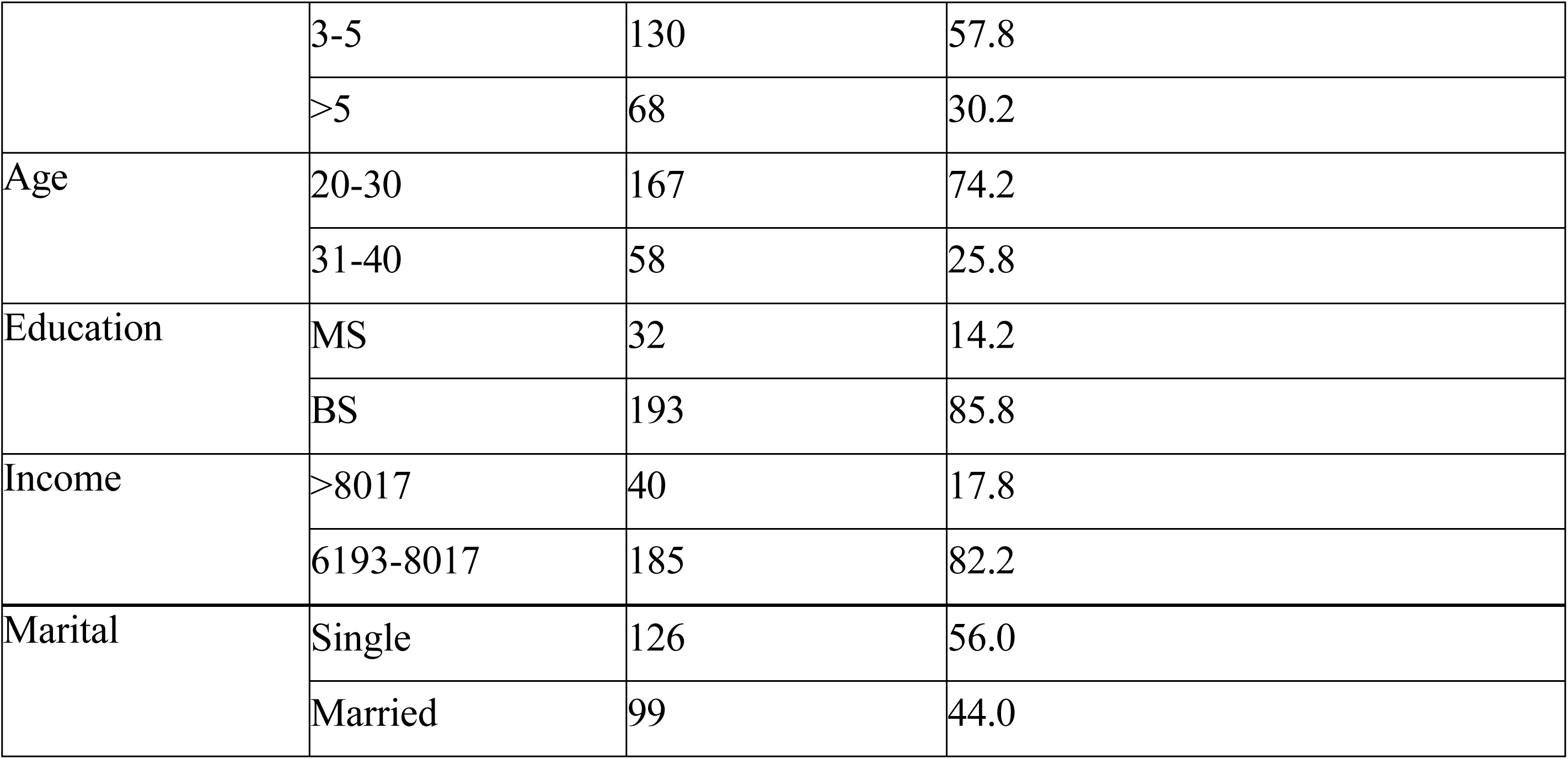
Socio-demographic characteristics of nurses in southern Ethiopia, selected hospitals, 2024(n=225)

#### Organizational factors

The majority of nurses (17.8%) and (89.3%) reported that there was a shortage of pressure-relieving devices (inadequate facility equipment) and a lack of guidelines for the risk assessment and preventative practices of PI, respectively. More than three-quarters of nurses (64.9%) of nurses reported that not satisfied with nursing leadership and 19.1% reported staff shortages. Moreover, 70.7% of nurses reported that the hospital had formal training on pup. However, 66.7% reported that the institutions are general hospital. (Table2).

**Table 2.** Organizational factors of nurses on pressure ulcer prevention in southern Ethiopia selected hospitals, 2024 (n=225)

| Organizational items |  | Frequency | Percent |
| --- | --- | --- | --- |
| Formal training on pup | no | 66 | 29.3 |
|  | yes | 159 | 70.7 |
| Guideline on pup | no | 24 | 10.7 |
|  | yes | 201 | 89.3 |
| Nursing leadership | not satisfied | 146 | 64.9 |
|  | satisfied | 79 | 35.1 |
| Staff shortage | disagree | 182 | 80.9 |
|  | agree | 43 | 19.1 |
| inadequate Facility<br>equipment | disagree | 185 | 82.2 |
|  | agree | 40 | 17.8 |
| Type of institution | general | 150 | 66.7 |
|  | teaching | 75 | 33.3 |

#### Clinical factors

The majority of the nurses 186 (82.7%) of were using the existing guidelines about pressure ulcer prevention practices and of them 125 (55.6%) had not received training. 180(80%) nurses complain over workload and 190 (84.4%) of nurses had unproportionate nurses-to-patient ratio in the ward, and 218 (96.6%) and 217 (96.4%) of staff and patients were cooperative with the health care service, respectively. Areas of practice of respondents;- 31.6% from surgical ward, 28.9% orthopedics, 24.4%, from ICU and, 15.1% from obstetric and gynecologic ward. (Table 3).

**Table 3.** Clinical factors on pup of nurses on pressure ulcer prevention in southern Ethiopia selected hospitals, 2024 (n=225)

|  |  | Frequency | Percent |
| --- | --- | --- | --- |
| Received training on pup | No | 125 | 55.6 |
|  | Yes | 100 | 44.4 |
| Use of pup guideline | no | 39 | 17.3 |
|  | yes | 186 | 82.7 |
| Staff workload | no | 45 | 20.0 |
|  | yes | 180 | 80.0 |
| Unproportionated. Patient to nurse ratio | no | 35 | 15.6 |
|  | yes | 190 | 84.4 |
| Staff cooperativeness | no | 7 | 3.1 |
|  | yes | 218 | 96.9 |
| Patient cooperativeness | no | 8 | 3.6 |
|  | yes | 217 | 96.4 |
| Area of practice | Surgical | 71 | 31.6 |
|  | Oby& gynecology | 34 | 15.1 |
|  | ICU | 55 | 24.4 |
|  | Orthopedics | 65 | 28.9 |

### 4.2 Pressure ulcer prevention practice

Of the respondents, 137 (60.9%) had good practice, while 88 (39.1%) of the respondents had a poor practice with the mean value of 7.85 ± 1.723 for the prevention of pressure ulcers (Figure 3).

**Figure 3.**
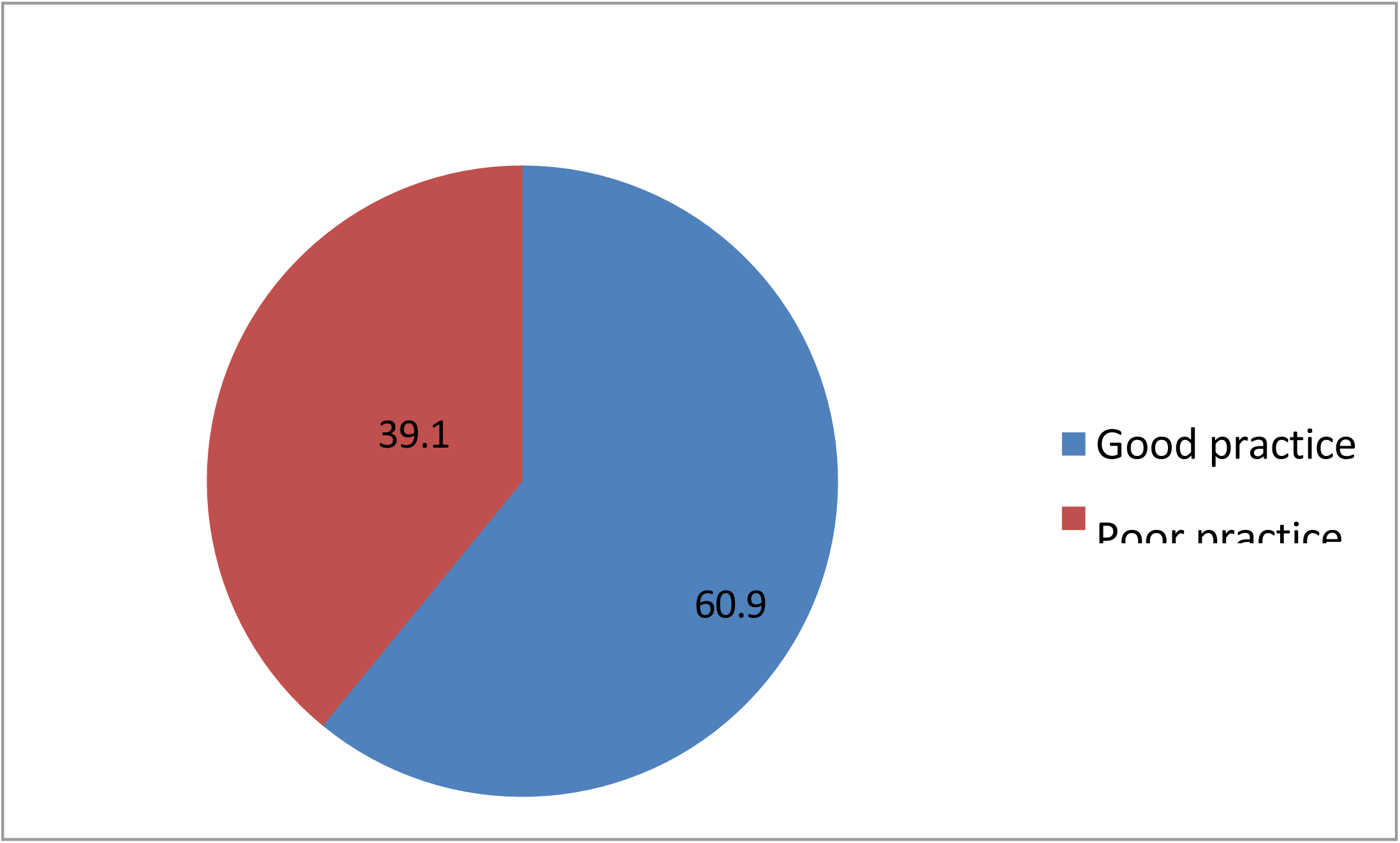
Pressure ulcer prevention practice among nurses working in southern Ethiopia hospitals, South Ethiopia, 2024 (n = 225).

**Fig 3.**
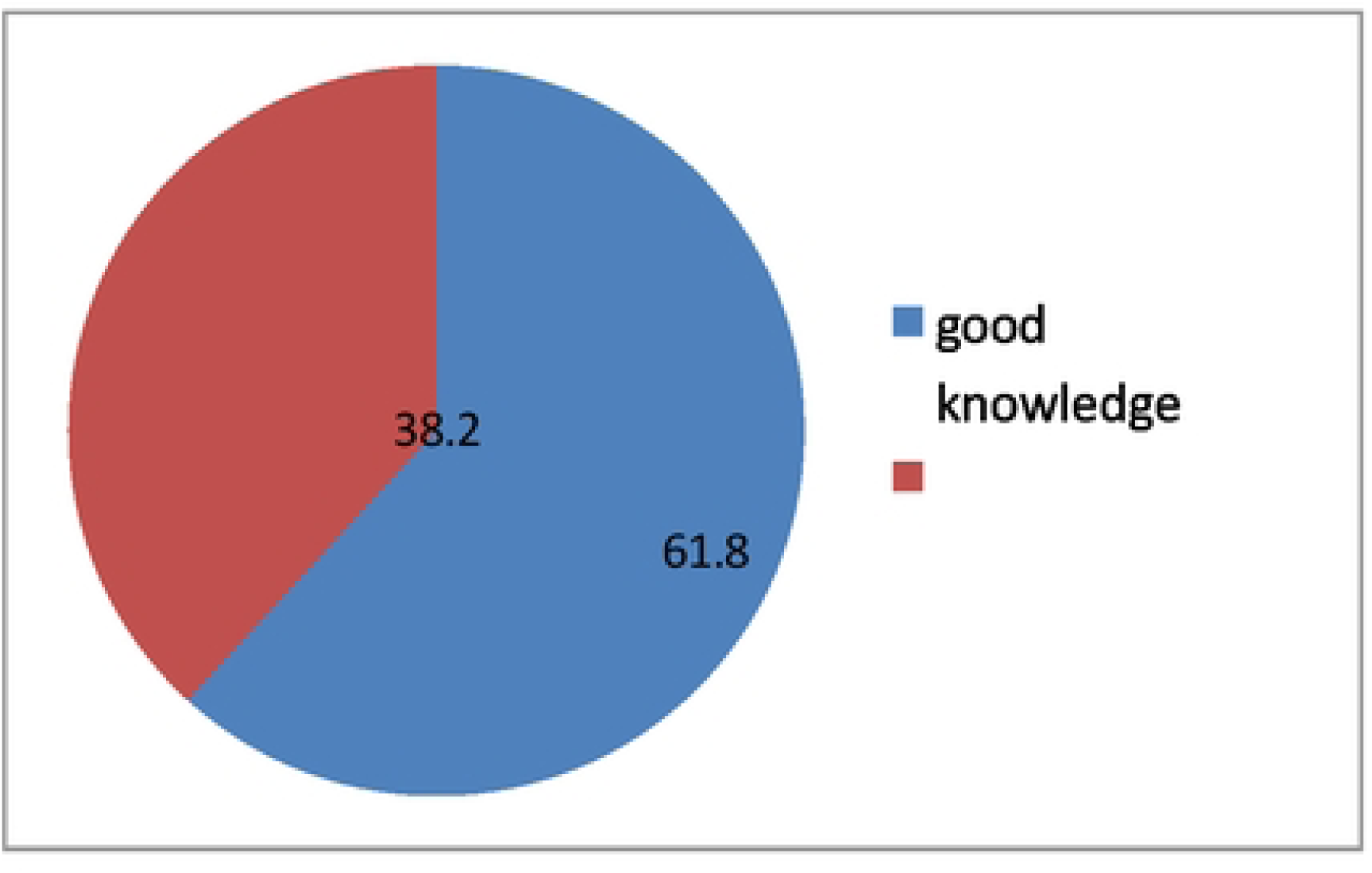
Pressure ulcer prevention knowledge among nurses working in southern Ethiopia hospitals, South Ethiopia, 2024 (n = 225).

### 4.3 Pressure ulcer prevention knowledge

Of the total respondents, 139 (61.8 %) of them had good pressure ulcer prevention knowledge; whereas the remaining 86 (38.2%) of the respondents had poor knowledge with a mean value of 18.04 ± 2.575 (Figure 4).

### 4.4 Factors associated with pressure ulcer prevention of practices

Knowledge, use of pressure ulcer prevention guideline, formal training on pressure ulcer prevention, and nursing leadership were associated with pressure ulcer prevention practices of nurses working in the hospital. Nurses having good knowledge of pressure ulcer prevention were 6.9 times more likely to have good pressure ulcer prevention practices than nurses who have a poor level of knowledge (AOR = 6.940 ; CI: 3.527-13.656).

Nurses using pressure ulcer prevention guidelines were 3.3 times having good practices about pressure ulcer prevention than nurses who did not use the guideline (AOR = 3.3574; CI: (1.317- 8.539).

Nurses those taken formal training on pressure ulcer prevention guideline was 2.2 times having good practices about pressure ulcer prevention than nurses who did not use the guideline (AOR = 2.253; CI: (1.044-4.862). Nurses who were satisfied with nursing leadership were 2.2 times more likely to have good practice than those who were not satisfied with nursing leadership AOR = 2.246; CI: (1.064-4.742). (Table 5).

### 4.5 Factors associated with pressure ulcer prevention of knowledge

Experience, marital status, use of pressure sore guideline and formal training and on pressure ulcer prevention were associated with pressure ulcer prevention knowledge of nurses working in the hospitals. Nurses with work experience of greater than five years were 3.5 times having good knowledge about pressure ulcer prevention than nurses work experience of one to two years(AOR = 3.52; CI: (1.343-9.244).

Nurses with work experience of three to five years were 2.9 times having good knowledge about pressure ulcer prevention than nurses work experience of one to two years (AOR = 2.995; CI: (1.302-6.887). Nurses using pressure ulcer prevention guidelines were 2.6 times having good knowledge about pressure ulcer prevention than nurses who did not use the guideline (AOR = 2.695; CI: (1.209-6.008).

The marital status of nurses being married was relatively 58% less likely to having good pressure ulcer prevention knowledge than being single (AOR = 0.414; CI: (0.219-0.780).

Nurses those taken formal training on pressure ulcer prevention guidelines were 2.9 times having good knowledge about pressure ulcer prevention than nurses who did not use the guideline (AOR = 2.968; CI :(1.508-5.843). (Table 4).

**Table 4.** Bivariate multivariate analysis of factors associated with knowledge on pressure ulcer prevention among nurses working in southern Ethiopia, 2023/2024and.

| Variables | Knowledge of nurses |  | COR (95% CI) | p-value | AOR (95%CI) | p-value |
| --- | --- | --- | --- | --- | --- | --- |
|  | Good % | Poor% |  |  |  |  |
| Experience |  |  |  |  |  |  |
| 1-2 | 15 | 21 | 1 |  |  |  |
| 3-5 | 87 | 43 | 2.83(1.33-6.04) | 0.007 | 2.99(1.30-6.88) | 0.010* |
| >5 | 37 | 21 | 2.47(1.05-5.78) | 0.038 | 3.52(1.34-9.24) | 0.011* |
| Marital status |  |  |  |  |  |  |
| Single | 87 | 39 | 1 |  |  |  |
| Married | 52 | 47 | 0.49(0.28-0.86) | 0.012 | 0.41(0.22-0.78) | 0.006* |
| Education |  |  |  |  |  |  |
| M.Sc. |  |  | 1 |  |  |  |
| B.Sc. |  |  | 0.40(0.17-0.98) | 0.045 |  |  |
| Use of PS guideline |  |  |  |  |  |  |
| no | 14 | 25 | 1 |  |  |  |
| yes | 125 | 61 | 3.66(1.78-7.53) | 0.000 | 2.69(1.21-6.01) | 0.015 * |
| Formal training |  |  |  |  |  |  |
| no | 29 | 37 | 1 |  |  |  |
| yes | 110 | 49 | 2.86(1.58-5.17) | 0.000 | 2.97(1.51-5.84) | 0.002* |
| Type of institution |  |  |  |  |  |  |
| general |  |  | 1 |  |  |  |
| Teaching |  |  | 0.54(0.31-0.95) | 0.34 |  |  |

□= significantly associated

## 5 Discussions

### 5.1 Pressure ulcer prevention knowledge and practices of nurses working in the hospitals

Pressure ulcer avoidance is a sign of high-quality treatment. The development and prevention of pressure ulcers are significantly impacted by nursing care. Thus, one important result that nurses should be aware of is pressure ulcers (Lyder and Ayello, 2005). So this study was aimed to assess nurse’s knowledge and practice on prevention of pressure ulcers and its associated factors in selected hospitals in southern Ethiopia.

In this study, 61.8 % of the participants were found to have good knowledge. This finding was similar to the studies reported in the Afghanistan(58.2,) (Muhammad *et al*., 2017), Brazil 63.4) (Souza Galvão *et al*., 2017), Nepal (65.66) (Hu, Sae-Sia and Kitrungrote, 2021), Australia(64.9) (Fulbrook, Lawrence and Miles, 2019), china 65.8% (Hu, Sae-Sia and Kitrungrote, 2021).The finding also resembles the studies from Dire Dawa 56.60 (Getie *et al*., 2020), Gurage (Tesfa Mengist *et al*., 2022) and Addis Ababa 67.3% (Dilie and Mengistu, 2015).

The nurse’s level of knowledge in this report was good (Figure 4) when relatively compared to the study findings reported from Slovakia (Halász *et al*., 2021), Saud Arabia (Guerrero *et al*., 2023), Saudi journal (Tullu, 2019), turkey (yilmazer, tüzer and erciyas, 2019), Uganda, (Mwebaza *et al*., 2014), UMgungundlovu District in South Africa, (Malinga and Dlungwane, 2020) show that nurses have poor knowledge of pressure ulcer prevention.

Similarly, the levels of knowledge were poor in the studies conducted in Federally Administered Hospitals in Addis Ababa (Dilie and Mengistu, 2015), Gondar University Hospital (Nuru *et al*., 2015), Hawassa University comprehensive specialized hospital (Muhammed *et al*., 2020b), Wolaita sodo university (Awoke *et al*., 2022), and public hospitals in Wollega (Ebi, Hirko and Mijena, 2019) relative to this study finding.

The discrepancy may also be caused by higher educational attainment of healthcare professionals (qualification), as well as better sources of information, scientific proof, and study findings.

According to this study, the levels of pressure ulcer prevention practices have been good, scored above the mean level (Figure 3). This finding was consistent with the study conducted in Bangladesh (Reza *et al*., 2020) Gurage, (Tesfa Mengist *et al*., 2022) and Addis Ababa (Dilie and Mengistu, 2015). Also, the study finding was similar to studies in South Africa (Malinga and Dlungwane, 2020), Kenya (Njau, Mwenda and Njoroge, 2018), Gondar University, (Nuru *et al*., 2015) Harare regional state and Dire Dawa (Getie *et al*., 2020).

While studies conducted in Sweden (Gunningberg *et al*., 2015), Uganda (Mwebaza *et al*., 2014), Afghanistan (Muhammad *et al*., 2017), Indian (Sawant and Shinde, 2017), Saud (Guerrero *et al*., 2023), Egypt (‘Nurses’ knowledge of prevention and management of pressure ulcer at a Health Insurance Hospital in Alexandria’, 2011), Tigray (Berihu *et al*., 2020),Addis Ababa (Werku Etafa, Ebi; Zeleke Argaw, Menji; Belachew Melese, 2017), Wolaita Sodo (Awoke *et al*., 2022) had poor pressure ulcer prevention practices relative to this study finding (Figure 2), which was good pressure ulcer prevention practices.

The difference may be attributed to nurses’ high level of expertise and growing body of scientific information, as well as their use of pressure ulcer prevention recommendations.

### 5.2 Factors associated with pressure ulcer prevention knowledge and practices of nurses

Nurses having good knowledge of pressure ulcer prevention were 6.9 times more likely to have good pressure ulcer prevention practices than nurses who have a poor level of knowledge (Table 5). This output was mimicked with the studies conducted in Bangladesh (Online *et al*., 2020), South Africa (Malinga and Dlungwane, 2020), Kenya (Njau, Mwenda and Njoroge, 2018), Borno State in Nigeria (Uba *et al*., 2015).

**Table 5.** Bivariate and multivariate analysis of factors associated with practice on pressure ulcer prevention among nurses working in southern Ethiopia, 2023/2024.

| Variables | Practice of H |  | COR (95% CI) | p-value | AOR (95%CI) | p-value |
| --- | --- | --- | --- | --- | --- | --- |
|  | Good % | Poor % |  |  |  |  |
|  |  |  | Practice of HP |  |  |  |
| Education |  |  |  |  |  |  |
| M.sc | 25 | 7 | 1 |  |  |  |
| B.sc | 112 | 81 | 0.38(0.16-0.94) | 0.036 |  |  |
| knowledge |  |  |  |  |  |  |
| poor | 25 | 61 | 1 | 1 |  |  |
| good | 112 | 27 | 10.12(5.41-18.95) | 0.000 | 6.94(3.53-13.67) | 0.000* |
| Income |  |  |  |  |  |  |
| >8017 | 29 | 11 | 1 |  |  |  |
| 6193-8017 | 108 | 77 | 0.53(0.25-1.13) | 0.101 |  |  |
| Use of PS guideline |  |  |  |  |  |  |
| no | 11 | 28 | 1 |  |  |  |
| yes | 126 | 60 | 5.35(2.45-11.46) | 0.000 | 3.36(1.32-8.54) | 0.011* |
| Staff workload |  |  |  |  |  |  |
| no | 23 | 22 | 1 |  |  |  |
| yes | 114 | 66 | 1.65(0.855-3.79) | 0.135 |  |  |
| Formal training |  |  |  |  |  |  |
| no | 26 | 40 | 1 |  |  |  |
| yes | 111 | 48 | 3.56(1.96-6.48) | 0.000 | 2.25(1.04-4.86) | 0.039* |
| Nursing leadership |  |  |  |  |  |  |
| Not satisfied | 76 | 70 | 1 |  |  |  |
| satisfied | 61 | 18 | 3.12(1.68-5.79) | 0.000 | 2.25(1.06-4.74) | 0.034* |

The result is also similar to the reports displayed in UMgungundlovu District in South Africa, (Tesfa Mengist *et al*., 2022) Addis Ababa (Dilie and Mengistu, 2015), Harari regional state Dire Dawa city administration hospital (Getie *et al*., 2020). Knowledge is the lowest level of learning outcome that would produce the highest level of learning outcome; practices or skills might be the possible reason. The reason might be due to the nurse having good knowledge and gaining enough skill during their clinical attachment.

Using pressure ulcer prevention guidelines was significantly associated with pressure ulcer prevention knowledge and practice of nurses, which was 2.6 times having a good knowledge level than nurses who were not using pressure ulcer prevention guidelines (Table 4). This result was similar to the studies reported from Australia (Fulbrook, Lawrence and Miles, 2019), German (Hoviattalab *et al*., 2014), South Africa (Malinga and Dlungwane, 2020) Embu in Kenya, (Njau, Mwenda and Njoroge, 2018) and Addis Ababa (Dilie and Mengistu, 2015).

The result is also similar to reports in Federal Hospitals in Addis Ababa (Werku Etafa, Ebi; Zeleke Argaw, Menji; Belachew Melese, 2017) and Wollega (Ebi, Hirko and Mijena, 2019), Ethiopia. The reason might be due to the use of guidelines used as a direct source of information and hold sets of systematic procedures that were expected from nurses for pressure ulcer prevention and having good knowledge.

Respondents who were satisfied with the nursing leadership had good practice as compared to those who were not. This result was similar to the study displayed in Gondar northern Ethiopia (Nuru *et al*., 2015). Possible reason for this result might be nurses who are satisfied with the nursing leadership are happier on their working environment, so that they are motivated to invest all their knowledge, skills and experience on practices related to prevention of pressure ulcer.

Nurses who received formal training on pressure ulcer prevention were 2.6 times having a good knowledge, and practice level than who did not receive formal training on pressure ulcer prevention. Similarly in studies conducted in Swedish, (Gunningberg *et al*., 2015) and university of Gondar in northern Ethiopia (Nuru *et al*., 2015); revealed that nurses who had training were more knowledgeable than those who did not. This might be due to the fact that training increases the possibility of the trainees to get up to date information about pressure ulcer related preventions.

Nurses with work experience of greater than five years were 3.5 times having good knowledge about pressure ulcer prevention than nurses work experience of one to two years.

Similar finding was reported in study conducted in Nigeria (Ikpi, Etita and Owan, 2019); where years of experience were significantly associated with clinical practice and knowledge and Gondar University in northern Ethiopia (Nuru *et al*., 2015).

Other study done in Spain on Nurses’ knowledge and clinical practice of pressure ulcer care revealed that, the greater the working experience the higher the knowledge gained (Pancorbo-hidalgo, Garcı and Lo, 2007). The reason might be nurses with more years of working experience have more exposure to work with different professionals in their clinical service so that they can learn from their coworker’s experience. Also since they have more prolonged exposure to patient care, they have greater probability to become skilled at how to prevent pressure ulcer even from their own mistakes as compared to those who have less years of working experience.

This study found that nurses being married were significantly associated with pressure ulcer prevention knowledge, with relatively 58% less likely to having good pressure ulcer prevention knowledge than being single (Table 4). There were no similar studies with this finding. Being married is more associated with the knowledge of nurses than nurses being single, this difference may be caused by social challenges in day-to-day activities and spending more time with children, which can hinder or interfere with married nurse practitioners’ clinical practices and workplace knowledge.

## Conclusion and Recommendation

The overall level of nurses’ knowledge and practice was good. Variables such as having a good knowledge, use of the guideline, formal training on pressure ulcer prevention, and nursing leadership were significant factors for the practice of nurses, and marital status, work experience, use of the guideline, and formal training were statistically significant factors for pressure ulcer prevention knowledge. In order to improve nurses’ knowledge and practices regarding pressure ulcer prevention, health managers should provide capacity building through ongoing professional development and create and disseminate updated guidelines to hospitals. The study might have faced some limitations; such as nature of the study design, which cannot establish a temporal relationship between the predictors and the outcome variable.

**Recommendation; -** Based on the findings of this study following recommendations were given:

### For Health Professionals

The health professionals who work in inpatient department ought to keep up their pressure ulcer prevention practice and knowledge with updated guidelines.

Married health professionals should updated their knowledge of pressure sore prevention as hospital guidelines to improve knowledge in pressure ulcer prevention.

### For Hospital Administrators and Regional Health Bureaus

The health managers should continue giving capacity building through continuous professional development and develop and distribute updated guidelines to the hospitals, which would enhance the pressure ulcer prevention knowledge and practices of nurses.

In order to improve nurse’s knowledge in pressure ulcer prevention needs to be conducted by giving priority for married nurses is the primary points to enhance nurses’ knowledge about pressure ulcer prevention.

### For future researchers

Further research on using observational studies is needed to determine the actual rather than the perceived practice and knowledge to PU prevention.

## Data Availability

no

## ACKNOWLEDGMENT

I would like to extend my sincere gratitude to Hawassa University School of nursing for giving me this opportunity to conduct Thesis. I also want to express my deepest gratitude to Advisors for their support and who spent their precious time by providing constructive information in order to prepare this proposal.

The end, I want to express my heartfelt gratitude to those selected hospitals and participants for giving their valuable information.

## Reference

Awoke, N. e t al. (2022) ‘Pressure injury prevention practice and associated factors among nurses at Wolaita Sodo University Teaching and Referral Hospital, South Ethiopia: A cross-sectional study’, BMJ Open, 12(3).

Batiha, A.-M. (2018) ‘Critical Care Nurses’ Knowledge, Attitudes, and Perceived Barriers towards Pressure Injuries Prevention’, International Journal of Advanced Nursing Studies, 7(2), p. 117.

Bereded, D.T., Salih, M.H. and Abebe, A.E. (2018) ‘Prevalence and risk factors of pressure ulcer in hospitalized adult patients; A single center study from Ethiopia’, BMC Research Notes, 11(1), pp. 1–6.

Berihu, H. et al. (2020) ‘Practice on pressure ulcer prevention among nurses in selected public hospitals, Tigray, Ethiopia’, BMC Research Notes, 13(1), pp. 1–7.

Booth, S. et al. (2013) ‘Best Practice Statement’.

Dave, K. and Choudhary, R.D. (2020) ‘Effectiveness of a pressure ulcer prevention package (pupp) for patients admitted in intensive care units: An experimental study’, International Journal of Advances in Nursing Management, 8(4), pp. 273–278.

Davoudi-Kiakalayeh, A. et al. (2017) ‘Alloimmunization in thalassemia patients: New insight for healthcare’, International Journal of Preventive Medicine, 8, pp. 1–7.

Dilie, A. and Mengistu, D. (2015) ‘Assessment of Nurses’ Knowledge, Attitude, and Perceived Barriers to Expressed Pressure Ulcer Prevention Practice in Addis Ababa Government Hospitals, Addis Ababa, Ethiopia, 2015’, Advances in Nursing, 2015, pp. 1–11.

Ebi, W.E., Hirko, G.F. and Mijena, D.A. (2019) ‘Nurses’ knowledge to pressure ulcer prevention in public hospitals in Wollega : a cross-sectional study design’, pp. 1–12.

Espejo, E. et al. (2018) ‘Bacteremia associated with pressure ulcers: a prospective cohort study’, European Journal of Clinical Microbiology and Infectious Diseases, 37(5), pp. 969– 975.

Etafa, W. et al. (2018) ‘Nurses’ attitude and perceived barriers to pressure ulcer prevention’, BMC Nursing, 17(1), pp. 18–25

Fulbrook, P., Lawrence, P. and Miles, S. (2019) ‘Australian Nurses’ Knowledge of Pressure Injury Prevention and Management: A Cross-sectional Survey’, *Journal of Wound*, Ostomy and Continence Nursing, 46(2), pp. 106–112.

Gedamu, H. et al. (2021) ‘Level of nurses’ knowledge on pressure ulcer prevention: A systematic review and meta-analysis study in Ethiopia’, Heliyon, 7(7), p. e07648.

Gedamu, H., Hailu, M. and Amano, A. (2014) ‘Prevalence and Associated Factors of Pressure Ulcer among Hospitalized Patients at Felegehiwot Referral Hospital, Bahir Dar, Ethiopia’, Advances in Nursing, 2014, pp. 1–8.

Getie, A. et al. (2020) ‘Pressure ulcer prevention practices and associated factors among nurses in public hospitals of Harari regional state and Dire city administration, Eastern Ethiopia’, PLoS ONE, 15(12 December), pp. 1–14.

Guerrero, J.G. et al. (2023) ‘A Multicenter Assessment of Nurses’ Knowledge Regarding Pressure Ulcer Prevention in Intensive Care Units Utilizing the PUKAT 2.0’, SAGE Open Nursing, 9.

Gunningberg, L. et al. (2015) ‘Pressure ulcer knowledge of registered nurses, assistant nurses and student nurses: A descriptive, comparative multicentre study in Sweden’, International Wound Journal, 12(4), pp. 462–468.

Halász, B.G. et al. (2021) ‘Nurses’ knowledge and attitudes towards prevention of pressure ulcers’, International Journal of Environmental Research and Public Health, 18(4), pp. 1–9.

Hinojosa-caballero, D. (no date) ‘y tratamiento de las úlceras por presión and treatment of pressure ulcers’, pp. 182–188.

Hospital, B.R., Zone, N.S. and Birhan, D. (2022) ‘Prevalence and Associated Factors of Pressure Israel Shiferaw 1 *, Nigussie Ulcer among Hospitalized Patients in Debre Tadesse, Sileshi Shiferaw Description of the study Area’, (November), pp. 1–7.

Hoviattalab, K. et al. (2014) ‘Nursing practice in the prevention of pressure ulcers : an observational study of German Hospitals’, pp. 1513–1524.

Hu, L., Sae-Sia, W. and Kitrungrote, L. (2021) ‘Intensive care nurses’ knowledge, attitude, and practice of pressure injury prevention in china: A cross-sectional study’, Risk Management and Healthcare Policy, 14, pp. 4257–4267.

Ikpi, J., Etita, R. and Owan, M. (2019) ‘Knowledge And Practice Towards Prevention Of Pressure Ulcers Among Nurses In Selected Private Hospitals In Calabar Metropolis, Cross River State, Nigeria’, 8(1), pp. 1–08.

Khojastehfar, S., Najafi Ghezeljeh, T. and Haghani, S. (2020) ‘Factors related to knowledge, attitude, and practice of nurses in intensive care unit in the area of pressure ulcer prevention: A multicenter study’, Journal of Tissue Viability, 29(2), pp. 76–81.

Lachenbruch, C. et al. (2016) ‘Pressure ulcer risk in the incontinent patient analysis of incontinence and hospital-acquired pressure ulcers from the international pressure ulcer prevalence^TM^ survey’, *Journal of Wound*, Ostomy and Continence Nursing, 43(3), pp. 235– 241.

Lagourdette, C. et al. (2018) ‘INVOLVEMENT OF ADVANCED PRACTICE NURSE IN THE MANAGEMENT OF GERIATRIC CONDITIONS : EXAMPLES FROM DIFFERENT COUNTRIES’.

Li, J. et al. (2023) ‘Critical care nurses’ knowledge, attitudes, and practices of pressure injury prevention in China: A multicentric cross-sectional survey’, International Wound Journal, 20(2), pp. 381–390.

Lotfi, M. et al. (2019) ‘Iranian nurses’ knowledge, attitude and behaviour on skin care, prevention and management of pressure injury: A descriptive cross-sectional study’, Nursing Open, 6(4), pp. 1600–1605.

Lyder, C.H. and Ayello, E.A. (2005) ‘Chapter 12. Pressure Ulcers : A Patient Safety Issue’.

Malinga, S. and Dlungwane, T. (2020) ‘Nurses’ knowledge, attitudes and practices regarding pressure ulcer prevention in the umgungundlovu district, South Africa’, Africa Journal of Nursing and Midwifery, 22(2).

Muhammad, D. et al. (2017) ‘Knowledge, Attitude and Practices of Nurses Regarding Pressure Ulcers Prevention At a Tertiary Care Hospital of Peshawar, Khyber Pakhtunkhwa’, Cell, 333(2), p. 9482238.

Muhammed, E.M., et al. (2020a) ‘Nurses’ knowledge of pressure ulcer and its associated factors at Hawassa University comprehensive specialized hospital Hawassa, Ethiopia, 2018’, BMC Nursing, 19(1), pp. 1–8. Available at: 10.1186/s12912-020-00446-6.

Muhammed, E.M., et al. (2020b) ‘Nurses’ knowledge of pressure ulcer and its associated factors at Hawassa University comprehensive specialized hospital Hawassa, Ethiopia, 2018’, BMC Nursing, 19(1), pp. 1–15. Available at: 10.1186/s12912-020-00446-6.

Mwebaza, I. et al. (2014) ‘Nurses’ Knowledge, Practices, and Barriers in Care of Patients with Pressure Ulcers in a Ugandan Teaching Hospital’, Nursing Research and Practice, 2014, pp. 1– 6.

Njau, S.K., Mwenda, C.S. and Njoroge, G.K. (2018) ‘Practices by nurses to prevent pressure ulcers among patients at a level five hospital in Kenya’, (January). ‘Nurses’ knowledge of prevention and management of pressure ulcer at a Health Insurance Hospital in Alexandria’ (2011), pp. 262–268.

Nuru, N., et al. (2015) ‘Knowledge and practice of nurses towards prevention of pressure ulcer and associated factors in Gondar University Hospital, Northwest Ethiopia’, BMC Nursing, 14(1), pp. 1–8. Available at: 10.1186/s12912-015-0076-8.

Online, I.P. et al. (2020) ‘Asian Journal of Medical and Biological Research Nurses’ knowledge and practices regarding prevention and management of pressure ulcer for hospitalized patient’, 6(2), pp. 237–243.

Pancorbo-hidalgo, P.L., Garcı, F.P. and Lo, I.M. (2007) ‘Pressure ulcer care in Spain : nurses’ knowledge and clinical practice’.

Reza, H.M. et al. (2020) ‘Nurses’ knowledge and practices regarding prevention and management of pressure ulcer for hospitalized patient’, Asian Journal of Medical and Biological Research, 6(2), pp. 237–243.

Said, A. and Shidi, A. (2016) ‘PRESSURE ULCER MANAGEMENT IN OMAN : NURSES’ KNOWLED GE AND VIEWS Author’s Publications and Presentations:’, (September).

Sawant, N. and Shinde, M. (2017) ‘Nurses Knowledge and Practices towards Prevention of Pressure Ulcer in Tertiary Care Hospital’, 6(5), pp. 739–745.

Shrestha, A. (2018) ‘Nepalese Critical Care Nurses’ Competency Towards Pressure Ulcer Prevention’, 5(1).

Shrestha, N. and Shrestha, P. (2016) ‘Knowledge of Pressure Ulcer Management among Nurses’, 09(02), pp. 47–51.

Souza Galvão, N., et al. (2017) ‘Program in Nursing in Adult Health Knowledge of the nursing team on pressure ulcer prevention’, Rev Bras Enferm Rev Bras Enferm [Internet], 7070(22), pp. 294–300.

Tallier, P.C. et al. (2017) ‘Perioperative registered nurses knowledge, attitudes, behaviors, and barriers regarding pressure ulcer prevention in perioperative patients’, Applied Nursing Research, 36, pp. 106–110.

Tesfa Mengist, S., et al. (2022) ‘Pressure ulcer prevention knowledge, practices, and their associated factors among nurses in Gurage Zone Hospitals, South Ethiopia, 2021’, SAGE Open Medicine, 10.

Tirgari, B., Mirshekari, L. and Forouzi, M.A. (2018) ‘Pressure Injury Prevention: Knowledge and Attitudes of Iranian Intensive Care Nurses’, Advances in Skin and Wound Care, 31(4), pp. 1–8.

Tubaishat, A., Aljezawi, M. and Al Qadire, M. (2013) ‘Nurses’ attitudes and perceived barriers to pressure ulcer prevention in Jordan’, Journal of Wound Care, 22(9), pp. 490–497.

Tullu, M. (2019) ‘Writing the title and abstract for a research paper: Being concise, precise, and meticulous is the key’, Saudi Journal of Anaesthesia, 13(5), pp. S12–S17.

Uba, M. et al. (2015) ‘Knowledge, attitude and practice of nurses toward pressure ulcer prevention in University of Maiduguri’, Inernational Journal of Nursing and Midwifery, 7(4), pp. 54–60.

Werku Etafa, Ebi; Zeleke Argaw, Menji; Belachew Melese, H. (2017) ‘Nurses’ knowledge and Perceived Barriers about Pressure Ulcer Prevention for Admitted patients in Public Hospitals in Addis Ababa, Ethiopia’, Committee of Ten Policy Brief: African Development Bank, 2(August), pp. 1–6.

Wu, J. et al. (2022) ‘Nurses’ knowledge on pressure ulcer prevention: An updated systematic review and meta-analysis based on the Pressure Ulcer Knowledge Assessment Tool’, Frontiers in Public Health, 10, pp. 613–620.

Yilmazer, T., Tüzer, H. and Erciyas, A. (2019) ‘Knowledge and Attitudes Towards Prevention of Pressure Ulcer: Intensive Care Units Sample in Turkey’, Turkiye Klinikleri Journal of Nursing Sciences, 11(2), pp. 140–147.

Zhang, Y. Bin et al. (2021) ‘Knowledge, attitude, and practice of nurses in intensive care unit on preventing medical device–related pressure injury: A cross-sectional study in western China’, International Wound Journal, 18(6), pp. 777–786.

